# The longest persistence of viable SARS-CoV-2 with recurrence of viremia and relapsing symptomatic COVID-19 in an immunocompromised patient – a case study

**DOI:** 10.1101/2021.01.23.21249554

**Authors:** Chiara Sepulcri, Chiara Dentone, Malgorzata Mikulska, Bianca Bruzzone, Alessia Lai, Daniela Fenoglio, Federica Bozzano, Annalisa Bergna, Alessia Parodi, Tiziana Altosole, Emanuele Delfino, Giulia Bartalucci, Andrea Orsi, Antonio Di Biagio, Gianguglielmo Zehender, Filippo Ballerini, Stefano Bonora, Raffaele De Palma, Guido Silvestri, Andrea De Maria, Matteo Bassetti

**Affiliations:** Infectious Diseases Unit, Department of Health Sciences (DISSAL), University of Genoa, Genoa, Italy; Infectious Diseases Unit, Ospedale Policlinico San Martino, IRCCS for Oncology and Neurosciences, Genoa, Italy; Hygiene Unit, Ospedale Policlinico San Martino, IRCCS for Oncology and Neurosciences Genoa, Italy; Department of Biomedical and Clinical Sciences Luigi Sacco, University of Milan, Milan Italy; Center of Excellence for Biomedical Research, Cytofluorimetry Unit, University of Genoa; Biotherapy Unit, Ospedale Policlinico San Martino, IRCCS for Oncology and Neurosciences, Genoa, Italy; Clinic of Hematology, Department of Internal Medicine (DiMI), University of Genoa, Genoa, Italy; Hygiene Unit, Department of Health Sciences (DISSAL), University of Genoa, Genoa, Italy; Hygiene Unit, Department of Biomedical and Clinical Sciences Luigi Sacco, University of Milan, Milan Italy; Hematology Unit, Ospedale Policlinico San Martino, IRCCS for Oncology and Neurosciences Genoa, Italy; Infectious Diseases Unit, Ospedale Amedeo di Savoia, University of Turin, Turin, Italy; Immunology Unit, Ospedale Policlinico San Martino, IRCCS for Oncology and Neurosciences, Genoa, Italy; Department of Pathology and Laboratory Medicine, Emory University School of Medicine, Atlanta, USA; Division of Microbiology and Immunology Yerkes National Primate Research Center, Emory Vaccine Center, Atlanta, USA

**Keywords:** SARS-CoV-2, viral shedding, viral persistence, viremia, hematological, immunological response

## Abstract

**Background:** Immunocompromised patients show prolonged shedding of SARS-CoV-2 in nasopharyngeal swabs. We report a case of a prolonged persistence of viable SARS-CoV-2 associated with clinical relapses of COVID-19 in a lymphoma patient.

**Methods:** Nasopharyngeal swabs and blood samples were tested for SARS-CoV-2 by Real time-PCR (RT-PCR). On five positive nasopharyngeal swabs, we performed viral culture and next generation sequencing. We analysed the patients’ adaptive and innate immunity to characterize T and NK cell subsets.

**Findings:** SARS-CoV-2 RT-PCR on nasopharyngeal swabs samples remained positive with cycle threshold mean values of 22 ± 1·3 for over 8 months. All five performed viral cultures were positive and genomic analysis confirmed a persistent infection with the same strain. Viremia resulted positive in three out of four COVID-19 clinical relapses and cleared each time after remdesivir treatment. T and NK cells dynamic was different in aviremic and viremic samples and no SARS-CoV-2 specific antibodies were detected throughout the disease course.

**Interpretation:** In our patient, SARS-CoV-2 persisted with proven infectivity for over eight months. Viremia was associated with COVID-19 relapses and remdesivir treatment was effective in viremia clearance and symptoms remission, although it was unable to clear the virus from the upper respiratory airways. During the viremic phase, we observed a low frequency of terminal effector CD8+ T lymphocytes in peripheral blood that are probably recruited in inflammatory tissue for viral eradication. In addition we found a high level of NK cells repertoire perturbation with a relevant involvement during SARS-CoV-2 viremia.

**Funding:** None.

## Introduction

Persistence of SARS-CoV-2 viral shedding and recurrence of positive SARS-CoV-2 nasopharyngeal swab after a negative sample and symptoms remission have been documented both in asymptomatic and symptomatic patients.^1^ Nevertheless, viral infectivity cannot be determined with Real time-PCR (RT-PCR), which detects only viral RNA, and viral culture and genome sequencing to determine the infectivity and genomic asset of the virus cannot be routinely used in clinical practise, leading to uncertainty about definitions of re-infection, relapse, prolonged viral shedding and infectiousness. Studies assessing the infectivity of these isolates have documented that viral infectivity declines within ten days of infection, with a possible tail of another ten days, both in asymptomatic and symptomatic individuals.^2^ In immunocompromised patients, prolonged viral shedding with proven viral infectivity is increasingly been reported^3–7^ and in two cases it was associated with disease relapse.^3–4^

Adaptive and innate immunity play a vital role in viral clearance: during many acute infections, including SARS-CoV-2, the presence of CD8+ cytotoxic T cells capable of secreting an array of molecules such granzymes to eradicate virus from the host has been described.^8^ At the same time, CD4+ helper T cells can assist cytotoxic T cells and enhance their ability to clear pathogens. However, persistent stimulation by the virus may induce T cell exhaustion as a state of T cells dysfunction, leading to loss of cytokine production and reduced function.^9^

Our case represents the longest SARS-CoV-2 viral persistence with proven viral infectivity in an immunocompromised patient who experienced four clinically relevant COVID-19 relapses. Relapses were associated with positive viremia and responded to remdesivir treatment with viremia clearance and symptoms remission but with persistence of viable virus in the upper respiratory airways. We observed a difference in T and NK cells distribution between aviremic and viremic phases.

## Methods

RT-PCR on nasopharyngeal swabs was performed with Allplex 2019 nCOV Seegene – Three Target assay [sensitivity 98·2%, specificity 100%].^10^

Viral isolation and culture from nasopharyngeal swab samples was performed on VERO E6 cells (Figure 1). We extracted RNA for confirmation of SARS-CoV-2 by RT-PCR and next generation sequencing. We obtained full genome sequences from the supernatant at different time points with Miseq Illumina (Illumina, Inc. San Diego) using 2×150 paired-end sequencing. We mapped and aligned the results to the reference genome obtained from Gisaid (https://www.gisaid.org/, accession ID: EPI_ISL_412973)^11^ using Geneious software, v. 9.1.5 (http://www.geneious.com).^12^ We used Nextclade and Pangolin systems (freely available at https://clades.nextstrain.org/ and https://pangolin.cog-uk.io/, respectively)^13,14^ for strain classification.

**Figure 1.**
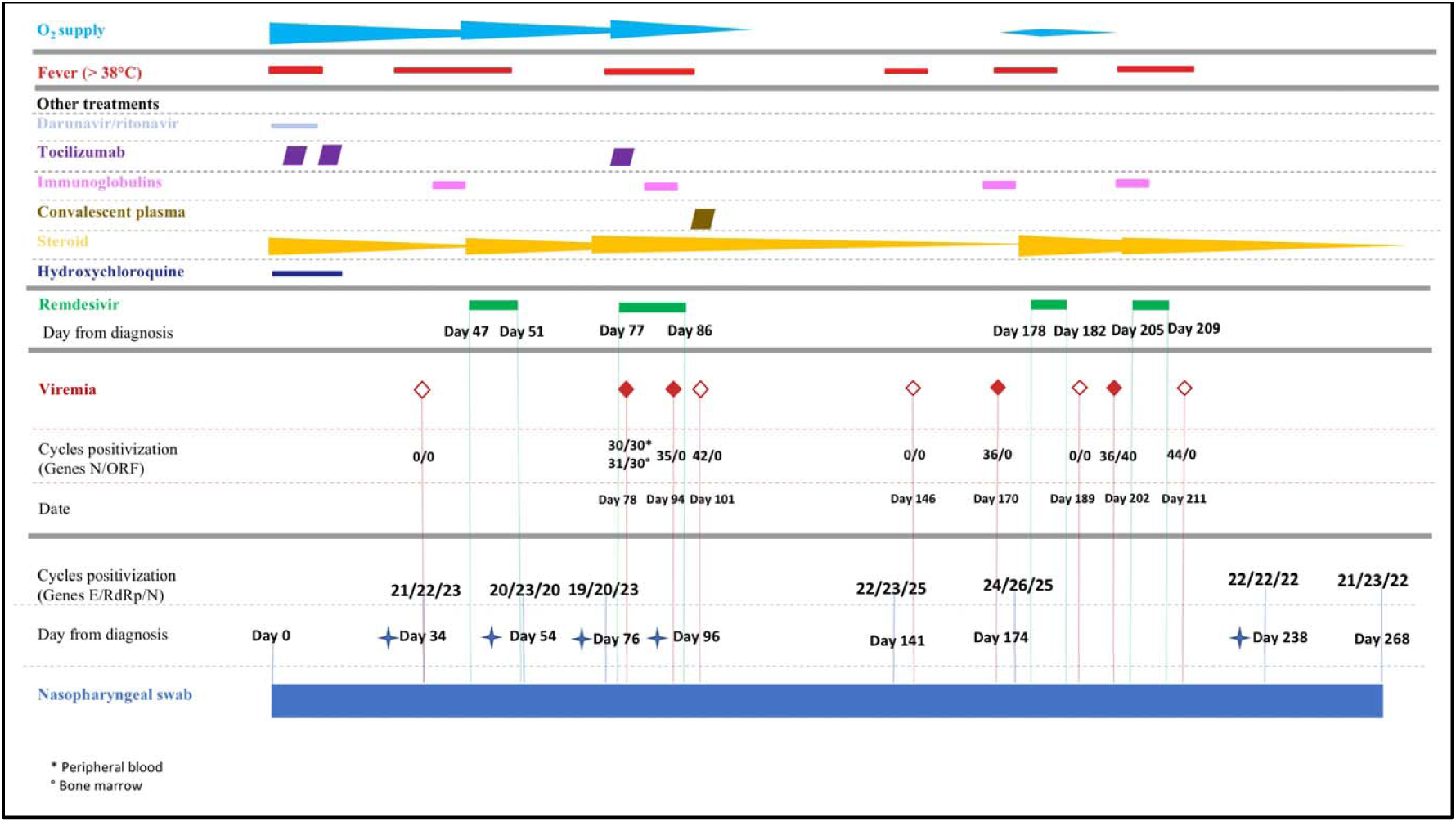
Timeline of SARS-CoV-2 RT-PCR Ct values in nasopharyngeal swabs, SARS-CoV-2 RT-PCR Ct values viremia [full rhombus = positive viremia, empty rhombus = negative viremia (Ct value for positive RT-PCR: 41 cycles)], administered treatments, O_2_ supply and fever from March 2020 to December 2020. The blue cross symbols indicate samples that underwent viral culture and sequencing.

SARS-CoV-2 IgG and IgM antibody testing was performed using Thermo Scientific− COVID-19 Total Antibody ELISA test.

We assessed T cells and NK cells activity on one viremic (Day 84) and one aviremic (Day 146) blood samples by BD Fortessa X20 flow cytometer (BD Biosciences) with the BD FACSDiva software (version 8.0; BD Biosciences) and by Activation Induced Marker (AIM) assay,^15^ a cytokine-independent approach using specific peptide megapools (MP), identified by bioinformatic approaches.^16^ We used multidimensional data reduction analysis through *t*-dependent Stochastic Neighbor Embedding (*t*-SNE) analysis to examine CD8+ T cells’ marker expression.

## Results

### Case history

During the SARS-CoV-2 initial wave in Italy,^17^ a 60-70 year-old male was hospitalized for the sudden onset of fever, dry cough and respiratory distress. The patient had been recently diagnosed with non-Hodgkin lymphoma (mantle cell type, blastoid variant stage IV), and had completed two cycles of R-BAC chemo-immuno-therapy (rituximab, bendamustine, cytarabine). A follow-up computed tomography (CT) documented lymphoma remission.

His nasopharyngeal swab resulted positive for SARS-CoV-2 Real-time PCR (RT-PCR) (Figure 1) and his CT scan showed bilateral interstitial pneumonia compatible with COVID-19. Blood tests showed anaemia (Hb 7·2 g/dL), absolute lymphopenia (0·20 x 10^9/L), increased CRP (28·5 mg/L cut-off 0·5 mg/L), IL-6 (57·2 ng/L, cut-off < 3·4 ng/L), ferritin (875 mcg/L, cut off 400 mcg/L), and D-dimer (900 mcg/L, cut off 500 mcg/L). We define his initial day of presentation with severe respiratory failure and fever and SARS-CoV-2 RT-PCR positive nasopharyngeal swab as day 0 of SARS-CoV-2 infection. The patient immediately required non-invasive mechanical ventilation with continuous positive airway pressure (CPAP). We started medical treatment with darunavir/ritonavir 800 mg/100 mg QD, hydroxychloroquine (400 mg BID), IV methylprednisolone (1 mg/kg/day), IV tocilizumab (8 mg/kg), and ceftaroline 600 mg BID according to the local COVID-19 treatment protocol in place at that time.

During the first month of hospitalization, the patient slowly recovered from acute respiratory failure; however, his follow-up SARS-CoV-2 RT-PCR nasopharyngeal swabs results were consistently positive. On day 38, he underwent immunoglobulin administration (0·4 mg/kg/die for 3 days). On day 45 his fevers returned, and a repeat lung CT demonstrated increased interstitial lesions with crazy-paving pattern. Bronchoalveolar fluid lavage results were as follows: SARS-CoV-2 RT-PCR positive; negative for bacterial, mycobacterial, and fungal growth with negative galactomannan; herpes simplex virus 1/2 DNA; positive for cytomegalovirus (CMV) DNA (3093 UI/ml). Serum CMV-DNA was 1181 UI/ml. Blood cultures, serum B-D glucan, and serum galactomannan were negative.

Several days after, the patient developed acute respiratory failure requiring ventilation with helmet CPAP. Given his clinical symptoms and CT findings, we concluded that his symptoms were consistent with COVID-19 relapse and started remdesivir (200 mg loading dose, then 100 mg QD) for a total of five days. We monitored his plasma concentration of remdesivir: T0 concentration at day 3 of 29·35 ng/ml for remdesivir and 81·82 ng/μl for its metabolite GS-441524. Peak values (2 hours after the end of administration) were, respectively 5364·31 ng/ml and 1310·86 ng/ml. After six days of helmet CPAP ventilation, the patient was progressively weaned to low-flow oxygen support (day 53). He was afebrile but had a positive repeat nasopharyngeal swab for SARS-CoV-2, with RT-PCR cycle threshold values comparable to those of his initial swabs (Figure 1), suggestive of viral persistence.

On day 66 he presented with a new onset of high fever (38.4 °C) and hypotension. Despite empiric broad-spectrum antibiotic therapy and successful treatment of an episode of CMV reactivation with gancyclovir, he remained persistently febrile. He developed severe respiratory failure again and required four days of ventilation with helmet CPAP. Repeat CT scan demonstrated persistent bilateral ground glass opacities and new bilateral pleural effusions. Repeat SARS-CoV-2 nasopharyngeal swab was positive, with RT-PCR cycle threshold (Ct) values comparable to previous assessments (Figure 1). SARS-CoV-2 viremia by RT-PCR was detected on day 78. A bone-marrow aspirate and trephine biopsy was performed which resulted negative for lymphoma; however, SARS-CoV-2 RNA viremia was detected by RT-PCR in blood from bone marrow aspirate. Having excluded other infections and lymphoma relapse, we concluded that SARS-CoV-2 was responsible and attributed his signs and symptoms to a second COVID-19 relapse. We treated the patient with a ten-day course of remdesivir and added a single tocilizumab infusion (8 mg/kg). On day 88 he underwent COVID-19 convalescent plasma infusion. The patient slowly recovered, with progressive weaning from oxygen therapy and clearance of SARS-CoV-2 viremia.

The third symptomatic COVID-19 relapse presented with persistent fever and new onset of mild respiratory failure requiring low-flow oxygen supplementation, followed by newly detected SARS-CoV-2 viremia (Figure 1). We initiated another five-day cycle of remdesivir with IV immunoglobulins (0·7 mg/kg/day for 3 days) on day 178. Viral clearance in blood was observed five days after the last dose of remdesivir. The patient was weaned from oxygen, became afebrile, and was subsequently discharged. Repeat CT scan and positron emission tomography excluded lymphoma relapse.

The fourth COVID-19 presented with fever in the absence of respiratory symptoms. SARS-CoV-2 viremia was detected (Figure 1) and we administered another five-day cycle of remdesivir (day 205 to 209), which resulted in viremia clearance. The patient was eventually discharged to a long-term facility for SARS-CoV-2 positive patients. His last nasopharyngeal swab specimen was collected on day 268 and resulted positive for SARS-CoV-2 (Figure 1). On day 271, the patient died following the onset of hematuria, hypotension, and general wasting.

### Viral sampling and analysis

RT-PCR Ct values of repeat nasopharyngeal swabs samples are displayed in Figure 1, together with viremia assessments. All nasopharyngeal swabs were positive for SARS-CoV-2, with Ct values persistently consistent with acute infection. We assessed viremia nine times during the disease course and it tested positive four times: all positive samples were obtained within the first from symptoms onset of three over four COVID-19 disease relapses with clinically relevant symptoms. Viremia clearance was observed all three times, always after remdesivir treatment (five, seven, and two days after the end of treatment, respectively).

We assessed viral infectivity by viral culture on five nasopharyngeal swab samples: cultures were all positive for viral replication with cytopathic effect observed at 48 hours. Comparing the isolated and sequenced viral strains by genomic analysis we observed an increasing number of in-host mutations from seven in April to 20 in November. We summarize the lineage assignment and observed mutations in Table 1. Patient sequences all belonged to clade 20B, corresponding to lineage B.1.1. According to literature data,^18–25^ no mutations related to possible resistance to remdesivir were observed, also as minority variant when considering a cut off of 1%.

**Table 1.** Lineage classification of strains included in the study according to Nextclade and Pangolin and identified mutations.

| Sequence Date | Clade | Pangolin | Total Aminoacid Changes | ORF1a | ORF1b | S | ORF3a | N | ORF14 |
| --- | --- | --- | --- | --- | --- | --- | --- | --- | --- |
| Day 34 | 20B | B.1.1 | 7 | P1640L | P314L | D614G |  | R203K,G204R | G50N |
| Day 54 | 20B | B.1.1 | 13 | K291E,K291N,T1322R,T1638I,R3561K,A4396V | P314L | D614G | S74F | R203K,G204R | G50N |
| Day 76 | 20B | B.1.1 | 15 | T1322R,T1638I,R3561K,A3705V,S4398L | P314L | D614G,W1214C,I1221K,G1223C | G44A | R203K,G204R | G50N |
| Day 96 | 20B | B.1.1 | 13 | T283P,K291T,K292T,V306I,T1322R,T1638I,S4398L | P314L | D614G |  | R203K,G204R | G50N |
| Day 238 | 20B | B.1.1 | 20 | R24C,V86F,T1322R,V1570L,T1638I,T4087I,S4398L | P314L,C455Y | H69Y,H69P,V70G,D614G,S982A | T151I | R203K,G204R | G50N |

### Immunological analysis

We performed SARS-CoV-2 serology repeatedly throughout the disease course. In all cases, no specific antibodies (IgG or IgM) were detected in the patient’s serum, even after infusion of convalescent plasma (which had an unspecified anti-SARS-CoV-2 titre).

Detailed analysis by flow cytometry on peripheral blood mononuclear cells to characterize T cell dynamic in viremic and aviremic samples showed that CD4+ and CD8+ T cells in viremic and aviremic samples clustered differently, highlighting profound modifications in both the regulatory and the effector arms of T cell adaptive immunity. We found decreased CD4 + / CD8 + ratio in both examined samples, with recovery of this ratio in the second (aviremic) sample (Figure 2, Panel 1A, B). With regard to regulatory T cells, their proportions were particularly high in the viremic phase, when the proportion of total CD4+ cells was lower, as shown by high frequency of CD4+FoxP3+CD25+ T cells, while their proportions dropped in the aviremic sample (Figure 2, Panel 2A, B).

**Figure 2.**
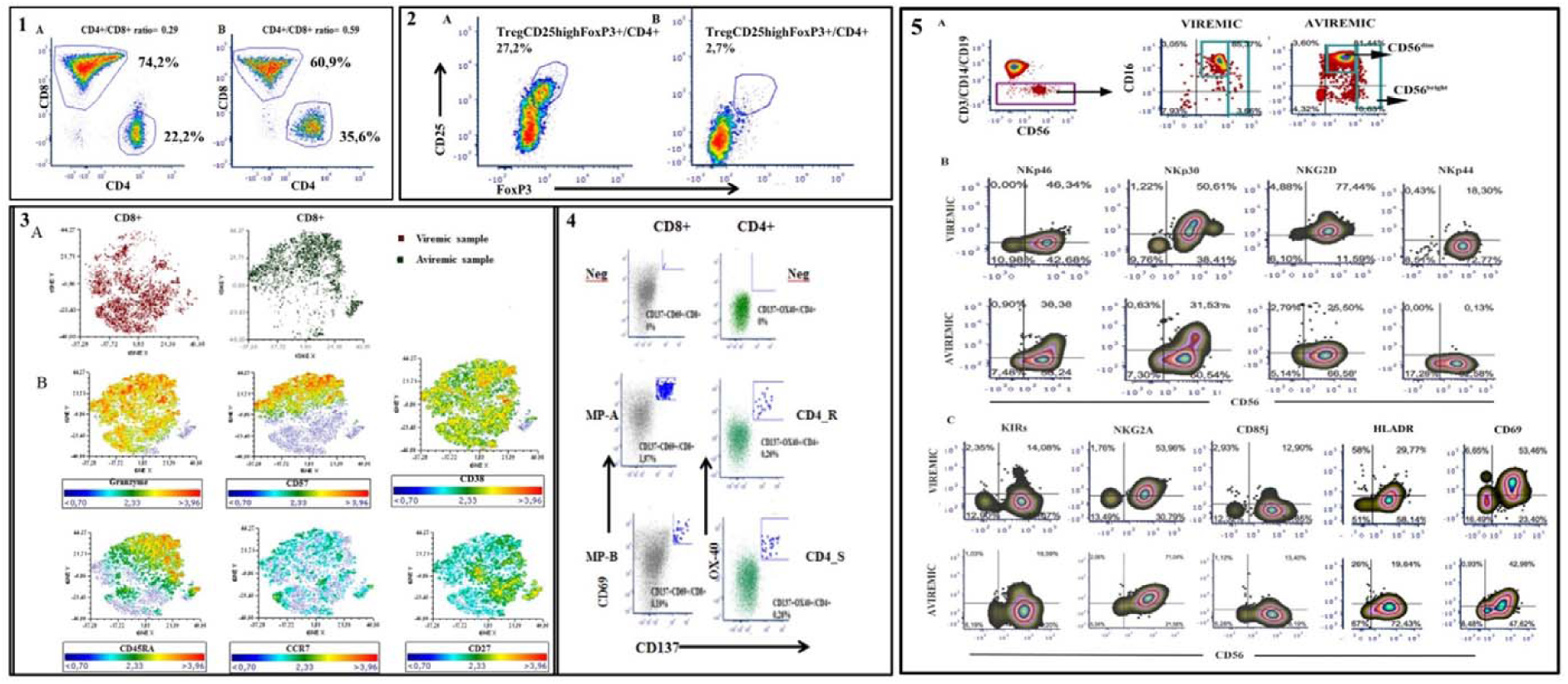
**Panel 1:** Comparison of CD4+/CD8+ ratio in CD3+T cells derived from viremic (panel 1A) and aviremic (panel 1B) samples. **Panel 2:** Comparison of CD4+FoxP3+CD25+ Treg cells in CD4+ T cells derived from viremic (panel 2A) and aviremic (panel 2B) samples. **Panel 3:** *t*-SNE analysis to the multiparametric analyses performed on CD8+ T cells in both samples. **Panel 3A:** t-SNE color plot of CD8+ lymphocytes in viremic and aviremic samples (panel A)**; Panel 3B :**t-SNE plots of concatenated CD8+ lymphocytes from viremic and aviremic samples according to the expression of CD45RA, CCR7, CD27, CD38, CD57 and Granzyme. **Panel 4** FACS plot analysis of SARS-CoV-2specific CD8 + and CD4+ T cell responses of aviremic sample. To measure SARS-CoV-2 specific CD4+ and CD8+ T cells, PBMCs have been stimulated with a spike MP (MP_S) and the class II MP representing all the proteome without spike (“non-spike”, MP CD4_R) and HLA A and HLA B peptide mega-pools (MP A, MP B). SARS-CoV-2 specific CD4+ T cell responses have been expressed as frequency of CD137+OX40+ cells on total CD4+ T cell population, whereas SARS-CoV-2 specific CD8+ T cell responses have been expressed as frequency of CD137+CD69+ cells on total CD8+ T cell population. **Panel 5** Flow cytometric analysis of NK cells at viremic or aviremic time points. **Panel 5A** Gating strategy to identify peripheral blood NK cells. **Panel 5B** Flow cytometric analysis of the expression of activating (NKp46, NKp30, NKp44, NKG2D) NK cell receptors. **Panel 5C** Flow cytometric analysis of inhibitory NK cell receptors (KIRs, NKG2A, CD85j) and activation markers (HLADR, CD69) expressed on the surface of NK cells circulating in peripheral blood in the presence or absence of viremia. *Note:* To identify SARS-CoV-2 specific CD8+ T cells, two class I peptide MPs have been used, based on epitope predictions for those 12 most common HLA A and B alleles, which collectively encompass 628 predicted HLA class I CD8+T cell epitopes from the entire SARS-CoV2 proteome (CD8 MP-A and MP-B).

SNE analysis performed on CD8+ T cells further highlighted marker expression differences between the two samples showing that CD8+ T cells clustered differently in viremic and aviremic samples (Figure 2, panel 3A). Accordingly, differences in phenotypic distributions were observed with regard to CD45RA, CCR7, CD27, CD38, CD57, and granzyme expression (Figure 2, panel 3B). In particular, the expression of CD57, a marker of terminally differentiated cells, was a highly discriminating factor when comparing the two samples. We observed a significant increase of CD57+ effector memory (CD45RA+CCR7-CD27-CD57+) in the aviremic sample that was associated with granzyme expression, confirming a relationship between cell differentiation and cytolytic enzymes in terminal effector CD8+ T lymphocytes. In fact, the dynamic picture of CD8+ T cell maturation showed a larger fraction of CD45RA+CCR7-terminal effector cells in aviremic sample (68% of total CD8+T cells) than that analysed in the viremic phase (18% of total CD8+T cells).

In the viremic phase a fraction of specific CD4+ and CD8+ T cells were described. In fact, results from AIM assay exhibited a significant frequency of CD137+CD69+ CD8+T cells specific for HLA A and HLA B peptide MPs (1·9% and 0·19% of total peripheral CD8+T, respectively) and a high percentage of CD137+OX40+ CD4+T cells specific for spike mega-pool (CD4-S) and all non-spike protein mega-pool (CD4-R) (0.26% of total peripheral CD4+T cells; Figure 2, panel 4).

Flow cytometric analysis of NK cells in PBMC, as defined by cells selected as CD3-CD14-CD19- by a negative gate, showed that during the clinical viremic phase associated to SARS-CoV-2 symptoms, CD56^bright^CD16^+/-^ NK cells were decreased compared to the subsequent clinical condition (Figure 2, panel 5A) characterized by absent symptoms following remdesivir treatment. In addition, the viremic phase was associated to a 3-fold increase in NKG2D+CD56+ and a 140-fold increase in NKp44+CD56+ NK cells, while minor or no increases were seen for the other natural cytotoxicity receptors (NKp30, NKp46) on circulating NK cells (Figure 2, panel 5B). Increased NKp44+ NK cell frequency during viremia was associated to increased NK cell activation as indicated by HLA-DR expression (Figure 2, panel C) accounting for 33% of CD56+NK cells. On the other hand, the circulation of NKp44+CD56+ NK cells was shut off when viremia was absent although some level of NK cell activation (21% of CD56+NK cells) persisted.

## Discussion

In this case study, we describe a case of 8-month persistence of SARS-CoV-2 RT-PCR positive nasopharyngeal swab in an immunosuppressed patient with non-Hodgkin lymphoma, with four clinically symptomatic relapses of COVID-19. During each relapse, the patient received a cycle of remdesivir that resulted in viremia clearance; however, viable virus remained detectable on nasopharyngeal swabs. To the best our knowledge, this is the longest persistence of SARS-CoV-2 RT-PCR positive nasopharyngeal swab to be reported. Our report suggests that this persistence may be due to viable SARS-CoV-2 virus that can determine a well-defined innate and adaptive cellular immune response compatible with disappearance of viremia, whose presence correlated with symptomatic relapses, as previously reported.^26,27^

Remdesivir was effective in clearing viremia and was associated with clinical recovery every time it was administered but was unable to clear SARS-CoV-2 from upper airways for yet unclear reasons that may include viral reservoir establishment or poor tissue PK/PD in this patient.

Since the disease course of this immunosuppressed patient was characterized by persistent viral shedding with intermittent viremia, we set to study whether an adaptive and innate immune response could be characterized in the different viremic phases. Taken together, the analyses highlighted profound differences in both the regulatory and the effector arm of T cell adaptive immunity in viremic and aviremic samples with an activated cytotoxic phenotype displayed by acute-phase SARS-CoV-2-specific T cells, as previously reported.^28,29^

During the viremic phase, we observed a high frequency of CD4+FoxP3+CD25+ T cells (T reg cells, tenfold higher than in the aviremic phase) and a reduced frequency of terminal effector CD8+ T lymphocytes (18% vs 68%) which is attributable to CD8+ T cells’ ability to migrate into tissues affected by inflammation to control the virus.

Furthemore, we observed a high level of NK cell repertoire perturbation with a relevant involvement of NKp44+ and NKG2D+ NK cells during SARS-CoV-2 viremia. This case report adds to existing literature on COVID-19 relapses and prolonged SARS-CoV-2 shedding in immunocompromised patients,^3–7^ highlighting the need for special attention in this particular population both in terms of clinical management comprising isolation measures and immunosuppressive treatment,^5,30^ and for future immunotherapeutic perspective, including vaccination.

## Data Availability

The authors confirm that the data supporting the findings of this study are available within the article and supplementary material.

## Authors contributions

CS, CD and MM drafted the manuscript and collected the data; BB, AO, AL, AB, GZ performed the virological analysis; ADM, FB, DF, AP, TA, RDP and CD carried out the immunological analysis and data interpretation; SB performed the therapeutic drug monitoring analysis; ADB, MB, GS, ED, FB, GB revised and provided conceptual insight to the manuscript.

## Acknowledgements

Thanks to BC and his family, for enduring the hardest times with dignity and determination and for always trusting us.

Thanks to Dr. Rosanna Vagge for her tireless job and for being always available.

A special thanks to Dr. Koh, who provided a fast and accurate English revision.

